# Pre-Diagnosis Dietary Patterns and Risk of Multiple Myeloma in the NIH-AARP Diet and Health Study

**DOI:** 10.1101/2023.09.20.23295639

**Authors:** Francesca Castro, Richa Parikh, Jelyn C. Eustaquio, Andriy Derkach, Janine M. Joseph, Alexander M. Lesokhin, Saad Z. Usmani, Urvi A. Shah

## Abstract

**Background:** Despite patient interest in knowing whether diet is linked to multiple myeloma (MM), there is limited research on dietary patterns and MM risk. Two studies have assessed this risk, albeit with a small number of MM cases. The EPIC-Oxford cohort and Oxford Vegetarian study (65 MM cases) showed that fish eaters, vegetarians and vegans had significantly reduced MM risk compared to meat eaters. The Nurses’ Health Study and Health Professionals Follow-up Study (478 MM cases) showed a significantly increased MM risk in men with Empirical Dietary Inflammatory Pattern.

**Methods:** The NIH-AARP Diet and Health study is a prospective cohort of 567,169 persons who completed a food frequency questionnaire in 1995-1996 and were followed until December 2011. Healthy Eating Index-2015 (HEI-2015), Healthy Diet Score (HDS), alternate Mediterranean Diet (aMED) and healthful Plant-based Diet Index (hPDI) scores were calculated using *a priori* defined methods and grouped into quartiles, with higher scores reflecting healthier eating patterns. We prospectively evaluated the association between pre-diagnosis dietary patterns and MM incidence in this cohort. Hazard ratios (HR) and 95% confidence intervals (95%CI) were estimated using multivariate Cox proportional hazards models adjusted for age at study entry, sex, race, body mass index, education, and total energy intake (by residual method). Sensitivity analysis was conducted to assess reverse causality by excluding MM cases diagnosed within one year of follow-up.

**Results:** Among 392,589 participants (after exclusions), a total of 1,366 MM cases (59% males; 92% non-Hispanic whites) were identified during the follow-up period. Analysis revealed a significant association between hPDI scores and reduced MM risk (highest vs lowest quartile, HR 0.85; 95%CI 0.73-1.0; p=0.043) (Table). In sensitivity analysis (1,302 MM cases), the association was no longer significant (HR 0.87; 95%CI 0.74-1.03; p 0.09) but trended in the same direction. This may be due to small sample size, given MM is a rare disease. HEI-2015, HDS and aMED scores were not associated with MM risk.

**Conclusions:** A healthful plant-based diet was associated with reduced MM risk in the NIH-AARP cohort. These results will help oncologists and patients make informed choices about their diet. To our knowledge, this is the largest epidemiologic study to date assessing pre-diagnosis dietary patterns and MM risk.

## Manuscript

The incidence of multiple myeloma (MM), the second most common hematologic malignancy, is rising globally [1]. Therefore, research efforts focused on identifying risk factors to reduce risk of MM are essential. Several risk factors have been identified, some that are non-modifiable (older age, male sex, African American race) and others that are modifiable (obesity, dietary patterns, diabetes mellitus). Additionally, amongst 421 patients with plasma cell disorders, 82% had questions about their diet and 57% reported their questions were not addressed by their oncologists. Among those who received dietary guidance from their oncologists, 94% attempted to follow it [2].

Despite this, there is limited data on dietary patterns and MM risk with few MM cases in each study. The EPIC-Oxford cohort and Oxford Vegetarian study (65 MM cases) showed vegetarians and vegans had 77% lower relative risk to develop MM than meat eaters [Relative risk (RR) 0.23, 95% confidence interval (95%CI) 0.09-0.59] [3]. The Nurses’ Health Study and Health Professionals Follow-up Study (478 MM cases) showed the Empirical Dietary Inflammatory Pattern had a 16% increased MM risk in men [Hazard Ratio (HR) 1.16, 95%CI 0.96-1.24] [4].

Therefore, we evaluated the association between pre-diagnosis dietary patterns and MM risk in the NIH-AARP Diet and Health Study, a large population-based, prospective cohort study. To our knowledge, this is also the largest epidemiologic study on diet and MM risk to date with 1 366 MM cases.

In 1995-1996, a food frequency questionnaire was mailed to 3.5 million American Association of Retired Persons (AARP) members who were aged 50–71 years, residents of six US states (California, Florida, Louisiana, New Jersey, North Carolina, and Pennsylvania) or two metropolitan areas (Atlanta and Detroit) [5]. A total of 566 398 satisfactorily completed questionnaires were returned between October 1995 and February 1997. Records were excluded from our analysis if they were completed in duplicate (n=179), questionnaire was completed by a proxy respondent (n=15 760) or proxy variable was not available or unknown (56 134), or if the participant died or moved out of the study area before returning the questionnaire (n=582), withdrew from the study (n=1), had a history of cancer as determined by self-report or confirmed from registry data (n=77 805), had a cancer diagnosis date prior to enrollment (n=1 564) and or had implausible energy intake (<800 or >4200 kcal/d for men and <600 or >3500 kcal/d for women) (n=20 967). After these exclusions, our final baseline cohort included 392 589 participants.

Study participants were followed from the date the questionnaire was received until first cancer diagnosis or until participant moved out of study area or died or follow-up period ended on December 31, 2011. Incident MM cases were identified through linkage to state cancer registries. Case ascertainment has been estimated to be about 90% complete [6]. MM cases were defined per International Classification of Diseases for Oncology, Third Edition code (ICD-O-3).

In this analysis, we used four dietary indices – Healthy Eating Index-2015 (HEI-2015), Healthy Diet Score (HDS), alternate Mediterranean Diet (aMED), and healthful Plant-based Diet Index (hPDI), to assess diet quality. Many of these scores have been inversely associated with cancer risk and cancer mortality. However, these pre-diagnosis scores have not been evaluated for MM risk. Using MyPyramid Equivalents Database and other variables, we were able to calculate each component and index scores for aMED and hPDI, while the HEI-2015 and HDS scores were previously calculated and provided in the database. These scores were grouped into quartiles. Higher scores imply higher adherence and a healthier diet.

The HEI-2015 is a measure of diet quality and overall alignment with the U.S. Dietary Guidelines for Americans and uses an energy-adjusted density approach for calorie adjustment [7]. There are nine adequacy components including total fruits, whole fruits, total vegetables, greens and beans, whole grains, dairy, total protein foods, seafood and plant proteins, and fatty acids. There are four moderation components including refined grains, sodium, added sugars, and saturated fats. The HEI score ranges from 0-100.

The HDS was adapted from the Healthy Diet Indicator which was based on World Health Organization recommendations [8]. It comprises of 12 components, saturated fatty acids, polyunsaturated fatty acids, protein, total carbohydrates, dietary fiber, fruit and vegetables, pulses and nuts, total non-milk extrinsic sugars, cholesterol, fish, red meat and meat products and calcium. Participants scored 0 or 1 based on their intake of each of these 12 items. The highest score is 11 [8].

The aMED score [9] was adapted from the traditional Mediterranean diet score [10] to assesses the conformity with the Mediterranean dietary pattern. The traditional Mediterranean Diet score was based on intake of nine components - vegetables, legumes, fruit and nuts, dairy, cereals, meat and meat products, fish, alcohol and the ratio of monounsaturated to saturated fat [10]. Intakes above the median received 1 point, all other intakes received 0 points. Meat and dairy product consumption less than median received 1 point. The aMED score excluded potato products from the vegetable group, separated fruit and nuts into 2 groups, eliminated the dairy group, included whole-grain products only, included only red and processed meats for the meat group and assigned alcohol intake between 5 and 15g/d for 1 point [9]. Scores ranged from 0 to 9.

The overall plant-based diet index (PDI) and hPDI were developed by splitting 18 dietary components into three groups healthful plant foods (vegetables, whole grains, fruits, legumes, nuts, vegetable oils, tea/coffee), unhealthful plant foods (fruit juices, refined grains, potatoes, sugar sweetened beverages, sweets/desserts), and animal foods [11]. We utilized the hPDI, in which the highest score was given to the highest quintile of intake for each healthful food group available, and the lowest score given to the highest quintile of intake for unhealthful plant foods and animal foods available.

HR and 95%CI for MM risk in highest versus lowest quartiles of dietary index scores were estimated using multivariate Cox proportional hazards models with person-years as the underlying time metric. The models were adjusted for age at study entry, sex, race (non-Hispanic Whites, non-Hispanic Blacks, other minority), body mass index (BMI) (<18.5, 18.5-24.9, 25-29.9, 30-34.9, ≥35), and education (less than high school, high school, post-high school training/college and unknown). Dietary variables were energy adjusted by residual method. Sensitivity analysis was conducted to assess reverse causality by excluding MM cases diagnosed within one year of follow-up.

Among 392,589 participants, 58.6% were males, 92.1% were non-Hispanic Whites, 34.4% had a normal BMI and 5.4% completed less than high school (Table 1). A total of 1 366 MM cases were identified during median follow-up period of 15.6 years. Multivariate analysis revealed a statistically significant inverse association between hPDI scores and MM risk (Q4 vs Q1, HR 0.85, 95%CI 0.73-1.0, p=0.043) (Table 2). There was no significant association between HEI-2015 (p=0.206), HDS (p=0.454), aMED (p=0.594) scores and MM risk. In a sensitivity analysis (1 302 MM cases), hPDI was associated with similar reduced MM risk statistical significance was not achieved (HR 0.87, 95%CI 0.74-1.03, p=0.09).

**Table 1:** Baseline characteristics of study cohort.

| Table 1: Baseline characteristics of study cohort |  |  |
| --- | --- | --- |
|  | Overall (N= 392,589) | Multiple Myeloma (N=1366) |
| **Age at Entry** |  |  |
| >60 | 229,032 (58%) | 952 (70%) |
| 45-60 | 163,557 (42%) | 414 (30%) |
| **Sex** |  |  |
| Male | 230,130 (59%) | 961 (70%) |
| Female | 162,459 (41%) | 405 (30%) |
| **Race** |  |  |
| Non-Hispanic White | 361,486 (92%) | 1,216 (89%) |
| Non-Hispanic Black | 13,714 (4%) | 94 (7%) |
| Hispanic/Asian/Pacific Islander/American Indian/Alaskan Native/Unknown | 17,389 (4%) | 56 (4%) |
| **Body Mass Index** |  |  |
| <18.5 | 3,935 (1%) | 11 (1%) |
| 18.5-25 | 134,997 (34%) | 420 (31%) |
| 25-30 | 16,3455 (42%) | 609 (45%) |
| 30-35 | 59,669 (15%) | 227 (17%) |
| >35 | 23,645 (6%) | 75 (5%) |
| **Education** |  |  |
| Less than high school | 21,231 (5%) | 74 (5%) |
| Completed high school | 75,189 (19%) | 248 (18%) |
| Post-high school training/ College/Postgraduate | 286,512 (73%) | 1,019 (75%) |
| Unknown | 9,657 (3%) | 25 (2%) |

**Table 2:** Cox multivariate analysis for dietary pattern scores in quartiles and MM risk.

|  |  |  |  |  |  |
| --- | --- | --- | --- | --- | --- |
| Dietary Score | Q1 | Q2 | Q3 | Q4 | p-value |
| HEI-2015 | 1 | 1.15 (0.99, 1.34) | 1.01 (0.87, 1.18) | 1.01 (0.87, 1.19) | 0.2055 |
| HDS | 1 | 0.95 (0.81, 1.10) | 0.88 (0.76, 1.03) | 0.92 (0.79, 1.07) | 0.4537 |
| aMED | 1 | 1.02 (0.89, 1.18) | 1.0 (0.87, 1.16) | 0.92 (0.79, 1.07) | 0.5942 |
| **hPDI** | 1 | 1.02 (0.88, 1.17) | 1.05 (0.9, 1.23) | **0.85 (0.73, 1.00)** | **0.0429\*** |
| Hazard Ratio (95% confidence interval)<br>*Statistically significant<br>MM: Multiple myeloma; HEI-2015: Healthy Eating Index-2015; HDS: Healthy Diet Score; aMED: alternate Mediterranean Diet; hPDI: healthful Plant-based Diet Index; Q: Quartile. |  |  |  |  |  |

In this large prospective analysis of the NIH-AARP Diet and Health Study, we found that higher hPDI scores were associated with a reduced MM risk. HEI-2015, HDS and aMED scores were not significantly associated with MM risk. Our findings confirm the observations from the EPIC-Oxford and Oxford-Vegetarian study [3]. The Nurses’ Health Study and Health Professionals Follow-up Study cohorts demonstrated 15-24% lower MM-specific mortality for presumed healthy pre-diagnosis plant-based dietary patterns [Alternate HEI (AHEI-2010), aMED, dietary approaches to stop hypertension (DASH) and Prudent)] [HR range 0.76-0.85 per 1-standard deviation (SD) increase in scores] and a 16-24% higher MM-specific mortality in unhealthy pre-diagnosis dietary patterns (Western and inflammatory/insulinemic dietary patterns) (HR range 1.16-1.24, per 1-SD increase in scores) [12]. These findings also suggest that healthy pre-diagnosis dietary habits may reduce risk of death once MM develops.

Plant-based diets are rich in fiber and phytochemicals and are associated with reduced cancer risk and improved survival in cancer through multiple mechanisms such as improved BMI, decreased inflammation, decreased insulin levels, decreased insulin-like growth factor 1 and improved stool microbiome [13]. As the role of gut dysbiosis is being increasingly recognized in the pathogenesis of MM, diet, the largest driver of microbiome composition can no longer be ignored [14]. Preliminary results of a pilot nutrition-based intervention study (NUTRIVENTION) in individuals with precursor plasma cell disorders and BMI ≥25 (NCT04920084) show that a plant-based diet is feasible and leads to improvement in biomarkers associated with progression to MM such as insulin resistance and gut microbiome composition [15]. A larger randomized study of a plant-based diet is currently enrolling (NCT05640843) [16].

The strengths of this study include the large sample size of MM cases and prospective nature of the study. A limitation of this study is the predominantly White population despite a higher MM incidence in Blacks. Further studies are warranted to better understand the underlying molecular and biological mechanisms.

In conclusion, the findings of this largest dietary epidemiologic study in MM are highly suggestive of a reduced MM risk with a healthful plant-based diet. These observations will help oncologists guide patients to make informed healthy dietary choices.

## Acknowledgements

The authors would like to thank the NIH AARP Diet and Health Study Team and Participants. This research was supported [in part] by the Intramural Research Program of the NIH, National Cancer Institute. Cancer incidence data from the Atlanta metropolitan area were collected by the Georgia Center for Cancer Statistics, Department of Epidemiology, Rollins School of Public Health, Emory University, Atlanta, Georgia. Cancer incidence data from California were collected by the California Cancer Registry, California Department of Public Health’s Cancer Surveillance and Research Branch, Sacramento, California. Cancer incidence data from the Detroit metropolitan area were collected by the Michigan Cancer Surveillance Program, Community Health Administration, Lansing, Michigan. The Florida cancer incidence data used in this report were collected by the Florida Cancer Data System (Miami, Florida) under contract with the Florida Department of Health, Tallahassee, Florida. The views expressed herein are solely those of the authors and do not necessarily reflect those of the FCDC or FDOH. Cancer incidence data from Louisiana were collected by the Louisiana Tumor Registry, Louisiana State University Health Sciences Center School of Public Health, New Orleans, Louisiana. Cancer incidence data from New Jersey were collected by the New Jersey State Cancer Registry, The Rutgers Cancer Institute of New Jersey, New Brunswick, New Jersey. Cancer incidence data from North Carolina were collected by the North Carolina Central Cancer Registry, Raleigh, North Carolina. Cancer incidence data from Pennsylvania were supplied by the Division of Health Statistics and Research, Pennsylvania Department of Health, Harrisburg, Pennsylvania. The Pennsylvania Department of Health specifically disclaims responsibility for any analyses, interpretations or conclusions. Cancer incidence data from Arizona were collected by the Arizona Cancer Registry, Division of Public Health Services, Arizona Department of Health Services, Phoenix, Arizona. Cancer incidence data from Texas were collected by the Texas Cancer Registry, Cancer Epidemiology and Surveillance Branch, Texas Department of State Health Services, Austin, Texas. Cancer incidence data from Nevada were collected by the Nevada Central Cancer Registry, Division of Public and Behavioral Health, State of Nevada Department of Health and Human Services, Carson City, Nevada. We are indebted to the participants in the NIH-AARP Diet and Health Study for their outstanding cooperation. We wish to acknowledge Dr. Arthur Schatzkin who was instrumental in conceiving and establishing the NIH-AARP Diet and Health Study. We also thank former and current study leaders at the National Cancer Institute and AARP, including Louise A. Brinton, Laurence S. Freedman, Albert R. Hollenbeck, Victor Kipnis, Michael F. Leitzmann, Linda M. Liao, Charles E. Matthews, Yikyung Park, Rashmi Sinha, Amy F. Subar and Mary H. Ward. Additionally, we are thankful to Sigurd Hermansen and Kerry Grace Morrissey from Westat for study outcomes ascertainment and management and Leslie Carroll and her team at Information Management Services for data support and analysis.

This study was funded by the Center for Hematologic Malignancies at MSKCC and in part through the NIH/NCI Cancer Center Support Grant P30 CA008748. Urvi A. Shah has received the American Society of Hematology Scholar Award, the NCI MSK Paul Calabresi K12 Career Development Award for Clinical Oncology K12CA184746, the MSK Parker Institute of Cancer Immunotherapy CDA and the International Myeloma Society CDA as well as additional funding support from HealthTree Foundation, the Allen Foundation Inc, the Paula and Rodger Riney Foundation, the Willow Foundation and the David Drelich, MD, CFP, Irrevocable Trust. U. A. Shah also received non-financial support from American Society of Hematology Clinical Research Training Institute and TREC Training Workshop R25CA203650 (PI: Melinda Irwin). A.M. Lesokhin received research support from NIH/NCI Cancer Center Support Grant P30CA008748, NCI 1R01CA249981–01, Sawiris Family Fund, and Paula and Rodger Riney Foundation.

## Author Contributions

Concept and design: RP, FC, AD, UAS. Acquiring data: RP, FC, AD, UAS. Analyzing data: RP, FC, AD, JE, JMJ, UAS. Drafting the manuscript: RP, FC, JE, UAS. Critical revision of the manuscript: FC, RP, JMJ, AML, SZU, UAS. Approving the final version of the paper: FC, RP, JE, AD, JMJ, AML, SZU and UAS. Submission: RP and UAS. RP and FC contributed equally to this work. All authors agreed to be accountable for all aspects of the work in ensuring that questions related to the accuracy or integrity of any part of the work are appropriately investigated and resolved.

## Data Availability

The data that support the findings of this study are available from the NIH-AARP Diet and Health Study, but restrictions apply to the availability of these data, which were used under license for the current study, and so are not publicly available. Data are however available from the authors upon reasonable request and with permission of the NIH-AARP Diet and Health Study team.

